# Multiomic approach and Mendelian randomization analysis identify causal associations between blood biomarkers and subcortical brain structure volumes

**DOI:** 10.1101/2023.03.30.23287968

**Authors:** Pritesh Jain, Madison Yates, Carlos Rubin de Celis, Petros Drineas, Neda Jahanshad, Paul Thompson, Peristera Paschou

## Abstract

Alterations in subcortical brain structure volumes have been found to be associated with several neurodegenerative and psychiatric disorders. At the same time, genome-wide association studies (GWAS) have identified numerous common variants associated with brain structure. In this study, we integrate these findings, aiming to identify proteins, metabolites, or microbes that have a putative causal association with subcortical brain structure volumes via a two-sample Mendelian randomization approach. This method uses genetic variants as instrument variables to identify potentially causal associations between an exposure and an outcome. The exposure data that we analyzed comprised genetic associations for 2,994 plasma proteins, 237 metabolites, and 103 microbial genera. The outcome data included GWAS data for seven subcortical brain structure volumes including accumbens, amygdala, caudate, hippocampus, pallidum, putamen, and thalamus. Eleven proteins and six metabolites were found to have a significant association with subcortical structure volumes. We found causal associations between amygdala volume and granzyme A as well as association between accumbens volume and plasma protease c1 inhibitor. Among metabolites, urate had the strongest association with thalamic volume. No significant associations were detected between the microbial genera and subcortical brain structure volumes. We also observed significant enrichment for biological processes such as proteolysis, regulation of the endoplasmic reticulum apoptotic signaling pathway, and negative regulation of DNA binding. Our findings provide insights to the mechanisms through which brain volumes may be affected in the pathogenesis of neurodevelopmental and psychiatric disorders and point to potential treatment targets for disorders that are associated with subcortical brain structure volumes.

## Introduction

Variations and dysfunctions of subcortical brain structures have been associated with numerous neurological and neuropsychiatric disorders such as Parkinson’s disease, different types of dementia, insomnia, schizophrenia, autism spectrum disorder (ASD), depression and post-traumatic stress disorder (PTSD) [1–7]. However, it is largely unknown how abnormalities of specific subcortical structures influence different traits and what processes may influence disease-related changes in subcortical brain structures. Understanding the relationship between brain volume and structure and neurological disease would help us better determine the underlying pathophysiological pathways. Such analysis could also be important in clinical practice, providing biomarkers that could be useful in disease diagnosis and patient management as well as helping to identify treatment targets for the various disorders associated with abnormalities in subcortical brain structure.

Recent large-scale multicenter studies such as the Enhancing Neuro Imaging Genetics through Meta-Analysis (ENIGMA) and UK Biobank (UKB) have put together neuroimaging and genomic data from tens of thousands of individuals and performed genome-wide association studies. This has led to the identification of genetic variants that are associated with subcortical brain structure volumes [8–10]. These studies have been followed by transcriptomic and epigenomic analysis to identify genes and epigenetic markers associated with regional brain volumes [11–13]. However, studies seeking to identify associations between regional brain volumes and other biomarkers such as proteins, metabolites and the microbiome are limited.

Here, we seek to address this gap, exploring the role of the proteome, metabolome, and microbiome in mediating brain structure changes which could lead to neurological disease. Proteins are the final product of gene expression and are an important intermediary phenotype that can provide insight into the cellular processes and functions that influence human biology and disease pathophysiology [14]. On the other hand, metabolites are small molecules that are a product and intermediates of cellular metabolism and play a pivotal role in cellular and physiological processes [15, 16]. The observed levels of such metabolites in biofluids can elucidate these processes. Finally, the human microbiota play an important role in the fermentation of non-digestible substrates as well as providing protection against foreign pathogens [17, 18]. A number of studies have found that changes in the level of different proteins, metabolites and the composition of the gut microbiome are associated with different metabolic, immunological as well as neurological disorders [19–22]. The importance of the level of different metabolites such as glucose, lactate and pyruvate in the cerebrospinal fluid (CSF) is well known and they are established biomarkers to study inflammation and malignancies in the brain [23]. The gut-brain axis (GBA) - which consists of bidirectional communication between the central and the enteric nervous system - is heavily influenced by the gut microbiota [24], establishing the importance of the microbiome in neurological functions and disorders.

Although the levels of these biomarkers in the body (especially metabolites and gut microbiome) are heavily influenced by environmental factors such as diet, medication and lifestyle [25–28], twin and family-based studies show that genetics also play an important role and they are highly heritable [29–31]. With advancements in profiling methods, large-scale studies can measure the levels of thousands of proteins and the various metabolites circulating in the blood and identify genetic variants which influence the level of these biomarkers [14, 32, 33]. Genome-wide association studies have also been performed to identify genetic variants that are associated with the composition of various bacterial taxa in the gut microbiome [34]. With results from these multi-omic studies at hand, there is the opportunity to investigate potential causal associations between such biological markers and subcortical brain structure volumes, using a two-sample Mendelian randomization (MR) approach.

MR analysis is a genetic epidemiological method that can help to determine putative causal associations between an exposure and an outcome using genetic variants as instrument variables [35, 36]. The method is conceptually similar to a randomized controlled trial which is based on the idea that the individuals receiving the treatment/drug (the instrument variable) are assigned randomly to the different groups [37]. Similarly, in MR studies, the SNPs are randomized by nature, assigned to offspring before birth and are not confounded by any environmental factor - thus satisfying the requirement of a randomized trial [36, 38]. This method is very powerful and can use the vast number of publicly available results of GWAS to identify causal associations between different exposures and outcomes. Indeed, studies undertaking this approach have identified causal associations between proteins and disorders such as depression, anorexia, ASD and many others [19, 39–41]. MR studies have also uncovered associations between the gut microbiome and autoimmune and cardiovascular disorders [42, 43]. MR studies for brain structures have also found causal associations between subcortical brain structure and neurological conditions like schizophrenia, anorexia, depression and other disorders[44–47]. However, so far no studies have examined associations between the different biomarkers and metrics of subcortical brain structures.

In this study, we sought to better understand the mechanisms and mediators that lead to the observed associations between brain structures and neurological and neuropsychiatric disease. In a systems biology approach, we integrated multi omic data with GWAS for subcortical brain volumes and employed a two-sample MR approach to ask if proteome, metabolome, and microbiome could be causally associated with volume of different subcortical brain structures. The central hypothesis of our study was that specific genetic variants influence subcortical brain volumes by altering levels of different biomarkers from the proteome, metabolome, or microbiome.

## Methods

### Study Design and Datasets

We applied a two-sample MR analysis to determine and identify causal associations between three multi-omic datasets (plasma proteome; metabolome; microbiome) and seven different subcortical brain structure volumes (accumbens, amygdala, caudate, hippocampus, pallidum, putamen and thalamus) using genetic variants as instrument variables. **Figure 1** shows the overall design of the analysis. The basic principle of MR is that SNPs (genetic instruments), which are significantly associated with modifiable exposure, would be causally associated with the exposure-related outcome. Three important assumptions are required for a valid genetic instrument and MR analysis. First, the instrument must be causally related to the exposure. Second, it must be independent of any confounders; and, finally, it should only be associated with the outcome through the exposure. In our current study, the genetic instruments for the different exposures were obtained from large-scale GWAS studies for each of the different omic datasets (information on these studies is shown in **Supplementary Table 1**). Overall, we obtained GWAS data on 2,994 plasma proteins, 237 blood metabolites and 103 microbial genera [32–34]. Our outcome dataset included the GWAS summary statistics for the seven subcortical brain structure volumes (adjusted for intracranial volume) obtained from the ENIGMA consortium [8].

**Figure 1:**
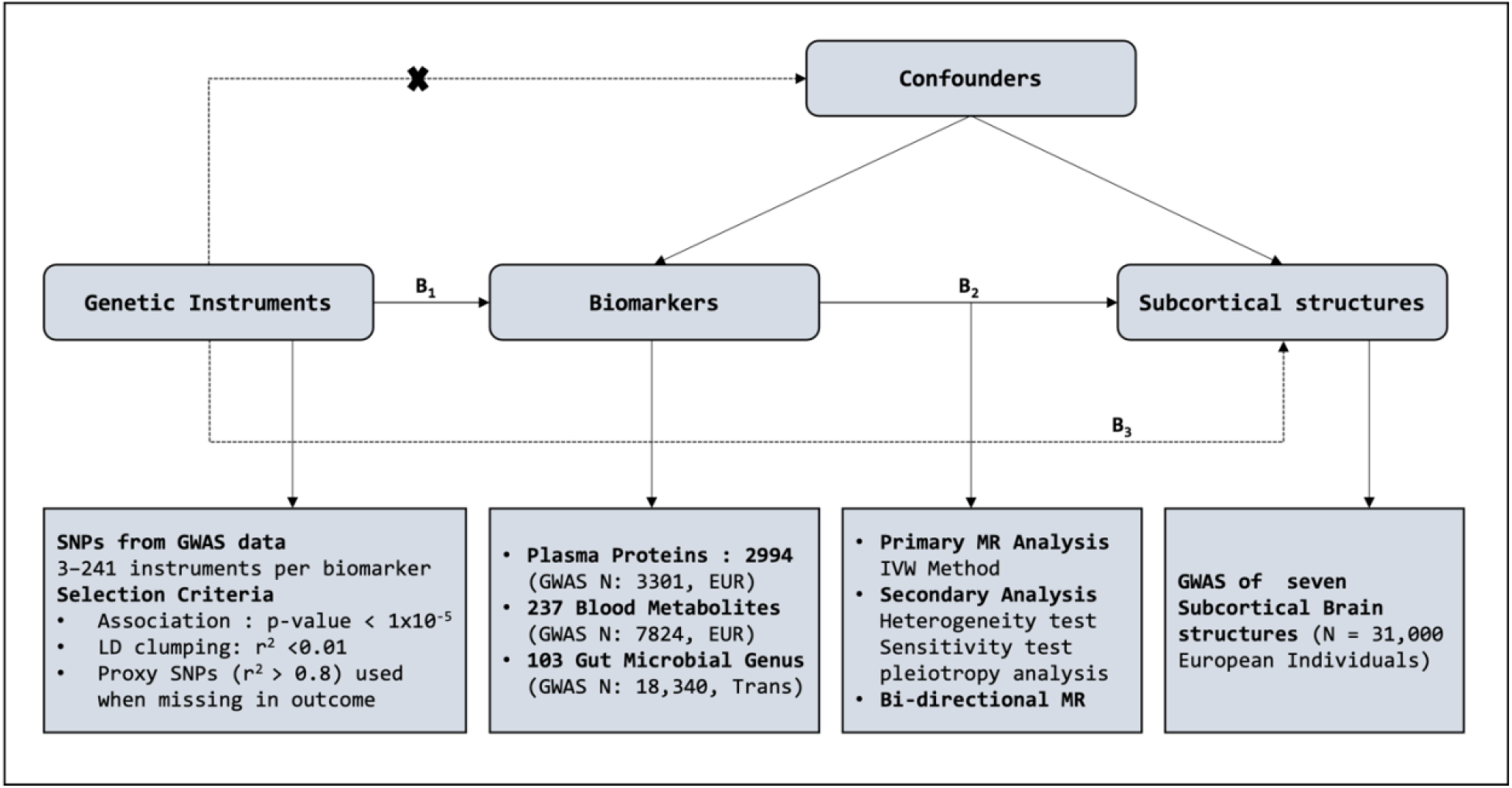
Study overview and design for MR analysis. SNP information for exposures and outcomes were extracted from GWAS summary statistics for each feature. B2 is the causal association of interest (Effect of Biomarkers on seven different subcortical brain structure volumes), estimated using B2 =B1/B3. B1 and B3 are the direct associations of the genetic variants on the exposure (biomarkers) and outcomes (subcortical structures) obtained from the GWAS studies. We also assume that the SNP instrument selected acts on the outcome only through exposure and not through any confounders. IVW: Inverse Variance Weighted.

### Selection of Genetic Instruments

The first step to performing MR analysis is the selection of instrument variables. We used a threshold of nominal significance (*P* < 1×10^−5^) to select SNPs from the GWAS summary statistics for each of the exposure variables. Ideally, genome-wide SNPs (*P* < 5×10^−8^) are used for MR analysis but a relatively relaxed threshold for the genetic instruments has been previously used in MR investigations when there were no or only a few genome wide SNPs available [40], [41, 48, 49]. To select independent SNPs, we performed LD clumping using PLINK2 with an *r*^*2*^ threshold of 0.01 using the 1000 Genomes European dataset as the reference panel [50]. The next steps of the analysis were performed using the TwoSampleMR package in R [51]. Once the independent SNPs were selected, we harmonized the exposure and outcome datasets to match the effect alleles, obtained the SNP effects and corresponding standard errors, and removed ambiguous SNPs with intermediate allele frequencies. In cases where a SNP was not available in the outcome dataset, a proxy SNPs with high LD with main SNP was used (LD at *r*^*2*^ > 0.8) for the analysis. We then evaluated the instrument strength of each of the exposures by estimating the proportion of variance explained by the SNPs (*R*^*2*^) and the *F*-statistic for each of the variables [52]. Typically, an *F* statistic >10 is considered sufficiently informative for MR analysis [53]We extracted a range of seven to 84 SNPs for the proteome data with an average R^2^ of 21% and the minimum *F* statistic was 20.56. The number of SNPs for the metabolites ranged from three to 241 with an average *R*^*2*^ of 13.1% and a minimum *F* statistic of 20.52. Finally, for the various microbial genera we extracted 3 to 22, with an average *R*^*2*^ of 3.2% and the lowest *F*-statistic of 20.46.The number of instrument variables, *R*^*2*^ and *F*-statistics for each individual biomarker is shown in Additional file 1.

### Two Sample MR Analysis and Statistical Validation

We used the inverse variance weighted (IVW) method of MR analysis to estimate the association between the different exposures and outcomes. The method provides a high power estimate and assumes that all the genetic instruments used for the analysis are valid. Significant associations of protein, metabolites and microbiomes with the different subcortical brain structures were identified after adjusting for multiple testing using the Benjamini-Hochberg false discovery rate (FDR) threshold of 0.05. We then performed downstream validation using other methods of MR estimation, heterogeneity analysis and pleiotropy analysis for the significant associations. Two methods - the weighted median method and MR-Egger method - were adopted as alternate methods to evaluate the robustness of causality and detect pleiotropy. These methods are useful to validate the results of the MR analysis in case we use SNPs that do not satisfy the assumptions for the analysis. The weighted median method provides a consistent estimate if less than 50% of the SNPs were invalid instruments[54] and the MR-Egger method was useful when up to 100% of the SNPs came from invalid instruments[55]. Cochran’s *Q* test was performed to test for heterogeneity, and pleiotropy was tested by performing an MR-Egger Intercept test and a leave-one out analysis. The directionality test to validate whether the genetic instruments were acting on the outcome through the exposure was tested using the MR Steiger directionality test, which calculates the variance explained in the exposure and the outcome by the instrumenting SNPs, and tests if the variance in the outcome is less than the exposure [56]. We also performed reverse MR analysis with the subcortical brain structure volume as exposure and the biomarkers as outcomes. This allows us to evaluate if there were any feedback loops between the brain structures and biomarker levels which could lead to false positive results. We used the same thresholds to select the genetic instruments from the GWAS studies of the subcortical structures and used the IVW method to estimate the association.

### Functional Enrichment Analysis

Functional enrichment analysis was performed using the gProfiler tool [57]. We tested for enrichment across different gene ontology terms, KEGG and reactome pathway databases, protein complexes and human phenotype ontology databases. A Bonferroni threshold was used to correct for multiple testing for all pathways tested. The pathway and enrichment analysis for metabolites was performed using the MetaboAnalyst platform [58].

## Results

### Investigating the Causal Association between Proteome and Subcortical Brain Structures

Using two sample MR analysis, we tested for potentially causal associations between 2,994 proteins and seven subcortical brain volumes (Additional file 2). Eleven proteins showed significant causal association with one of the subcortical brain structures as shown in **Figure 2** and **Supplementary Table 2**. Agouti Signaling Protein (ASIP) had the strongest association with putamen volume, with increase in the protein expression resulting in decrease in putamen volume (Beta: -28, *p-*value: 1.2×10^−8^). Plasma protease C1 inhibitor (SERPING1) and secretoglobin family 1C member 1 (SCGB1C1) were both found to be causally associated with accumbens volume, with the increase in expression of these proteins being associated with increase in the volume of accumbens (Beta: 6.3-9.7, *p*-value: 3×10^−5^ - 6.9×10^−7^). Increase in Granzyme A (GZMA) levels was found to be significantly associated with increase in amygdala volume (Beta: 17, p-value: 1.43×10*-5*). Two proteins had a significant causal association with caudate volume. Increase in Thioredoxin domain containing protein 12 (TXNDC12) levels was associated with increase in caudate volume (Beta: 11.7, *p*-value: 2.3×10^−6^), whereas Transmembrane protease serine 11D (TMPRSS11D) had a negative association (Beta: -26.8, *p*-value: 7.1×10^−7^). For the hippocampus, we found four proteins significantly associated and all of them had a negative association with volume of hippocampus. These included Copine-1 (CPNE1), Cardiotrophin-1 (CTF1), Selenoprotein S (VIMP) and and Protein CEI (C5orf38) (Beta: -21.2 to -25.9, *p*-value: 4.9×10^−5^ - 9.8×10^−7^). Finally, we found that increases in Chymotrypsinogen B (CTRB1) were significantly associated with decrease in the volume of thalamus (Beta: -23.9, *p*-value: 1.4×10^−5^). No proteins were found to be significantly associated with pallidum volume after multiple testing corrections.

**Figure 2:**
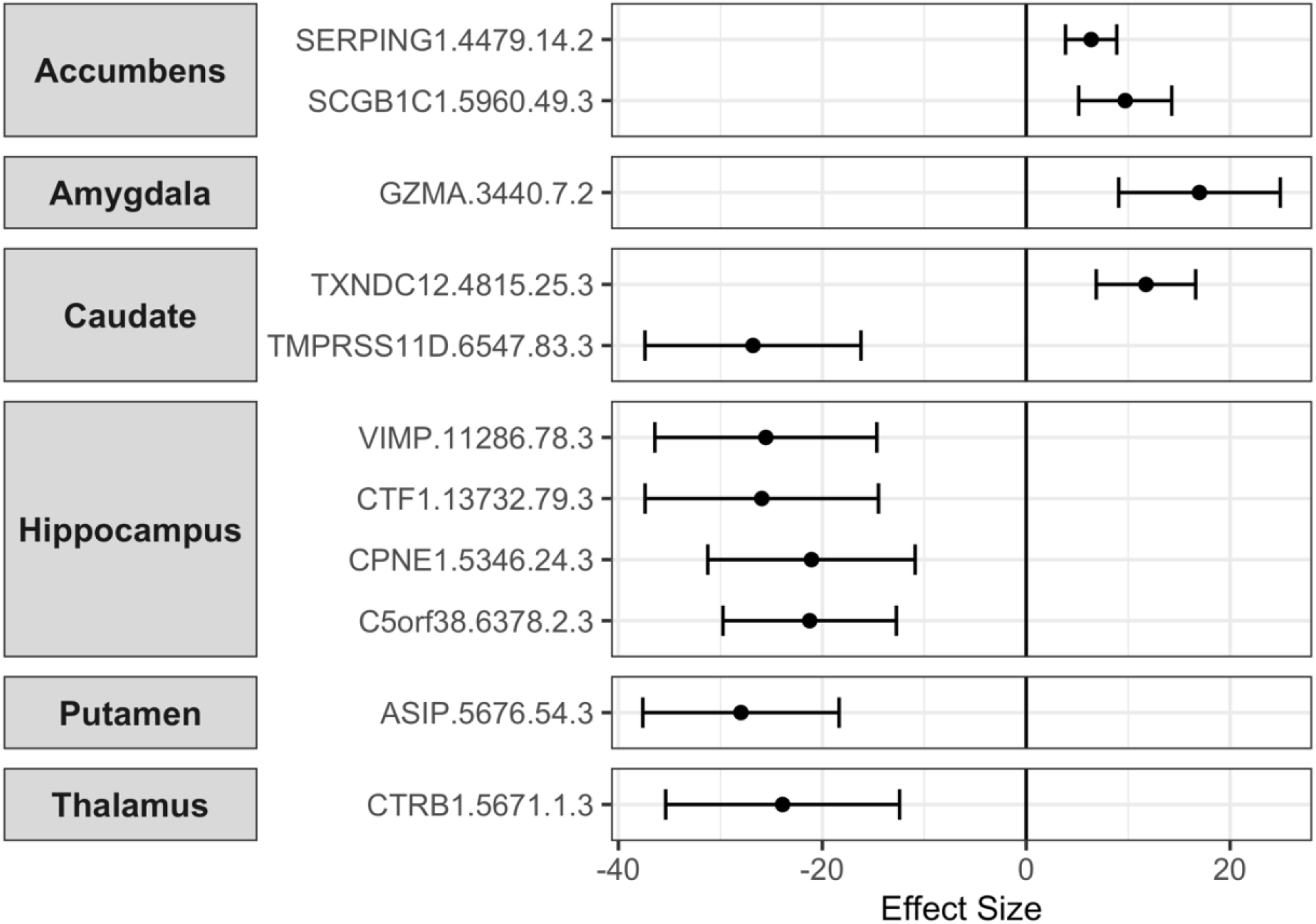
Significant causal associations between plasma proteins and subcortical brain structure volumes as uncovered via MR analysis. The Proteins were the exposures and the subcortical structures’ volume as outcomes. The associations were significant after FDR corrections for multiple testing.

### Investigating Causal Association between Metabolome and Subcortical Brain Structures

We proceeded to test for potentially causal association between metabolites and subcortical brain structure (Additional file 3). We found six metabolites to be significantly associated with one of the subcortical brain structure volumes (**Supplementary Table 3** and **Figure 3**). Among these, two metabolites had a causal association with amygdala volume. These included uridine levels which had a positive association (Beta: 255.9, *p*-value: 1.44×10^−4^) and Arachidonate which had a negative association with amygdala volume (Beta: -110.4, *p*-value: 2.54×10^−4^). We also found three metabolites significantly associated with thalamus volume which were Urate (Beta: -458.7, *p*-value: 3.7×10^−5^), 1-arachidonoylglycerphosphocholine (Beta: 269.7, *p*-value: 1.1×10^−4^) and N-acetylornithine (Beta: 72.4, *p*-value: 5.6×10^−4^). Increase in mannose levels was found to be causally associated with increase in caudate volume (Beta: 244.7, *p*-value: 5.5×10^−5^).

**Figure 3:**
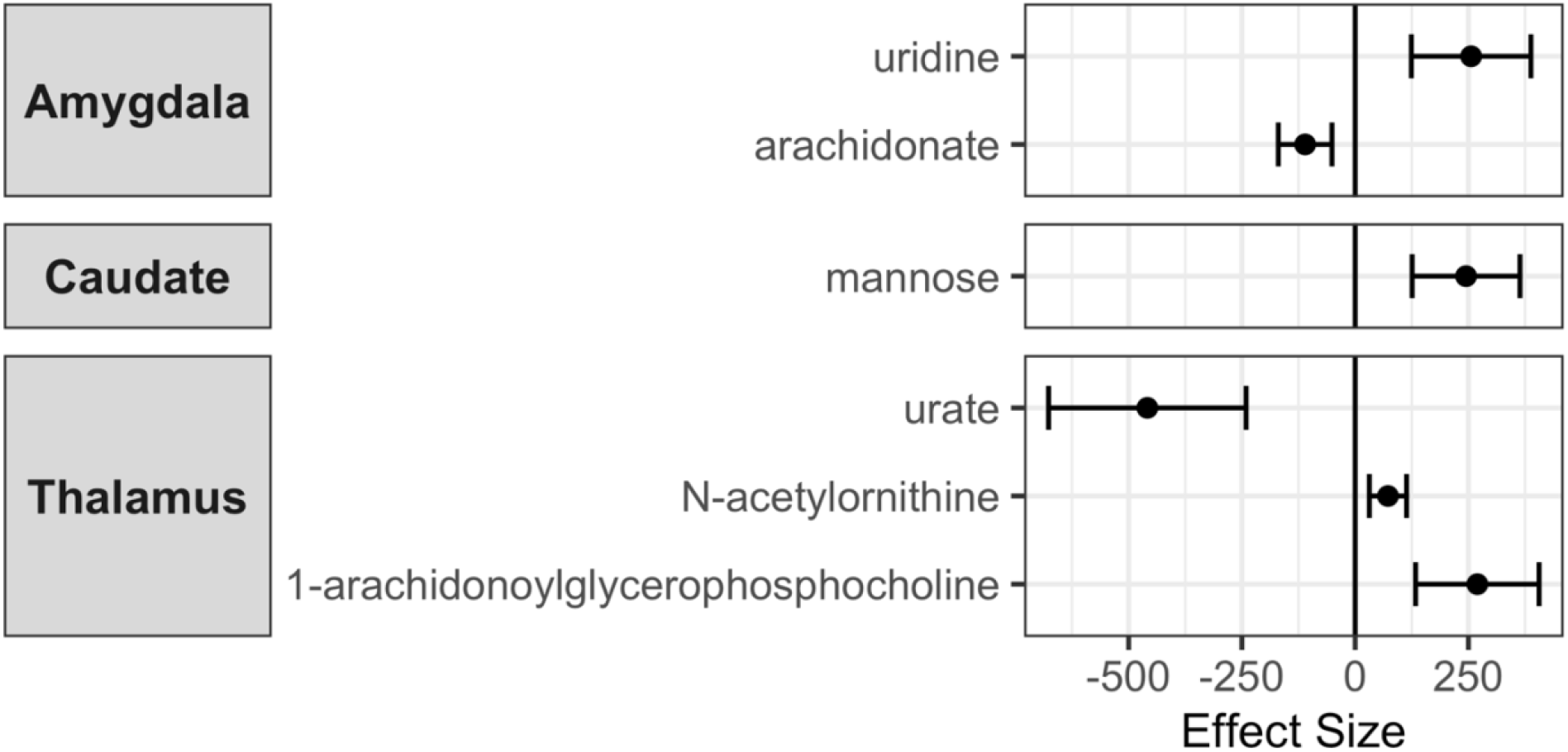
Significant causal associations between metabolites and subcortical brain structure volumes as uncovered via MR analysis. The metabolites were the exposures and the subcortical structures’ volume as outcomes. The associations were significant after FDR corrections for multiple testing.

### Investigating Causal Association between Microbiome and Subcortical Brain Structures

Here, we pursued MR analysis between 103 microbial genera as exposure and subcortical structure as outcome. Our analysis did not reveal any significant associations after multiple testing corrections (Additional file 4).

### Heterogeneity, Sensitivity and Pleiotropy Analyses

To determine the robustness and the validity of our results, we performed downstream statistical analysis to further increase the confidence in the observed associations. For all the significant associations identified in the primary analysis, we repeated the MR analysis using other methods such as the weighted median method and the MR-Egger method. We found that the associations were largely consistent with effects in the same direction and a significant *p*-value for the proteins (**Supplementary Table 4**). The MR-Egger estimate between the metabolites and subcortical brain volumes was found to be non-significant (**Supplementary Table 5**). We then determined if there was any heterogeneity in the genetic instruments used by calculating the Cochran’s *Q* statistic and found little to no evidence of heterogeneity (p-value: 0.094-0.99) for all proteins and metabolites (**Table 1A** and **1B**). Following this, we tested for pleiotropy of SNPs between exposure and outcome using the Egger intercept test and leave one out analysis. We found no evidence of pleiotropy (Egger Intercept *p*-value: 0.06-0.95) and leave one out analysis showed that removing any SNP did not greatly affect the association (**Table 1** and additional file 4). One of the assumptions of MR is that the instruments influence the exposure first and then the outcome through the exposure. To evaluate this, we used the MR-Steiger test which calculates the variance explained in the exposure and the outcome by the instrumenting SNPs, and tests if the variance in the outcome is less than the exposure. The test showed that for all the proteins and metabolites that had significant associations with subcortical volume, the variance of the genetic instruments in the exposure is always greater than the outcome - thus validating the assumption of MR (**Supplementary Tables 6 and 7**).

**Table 1:**
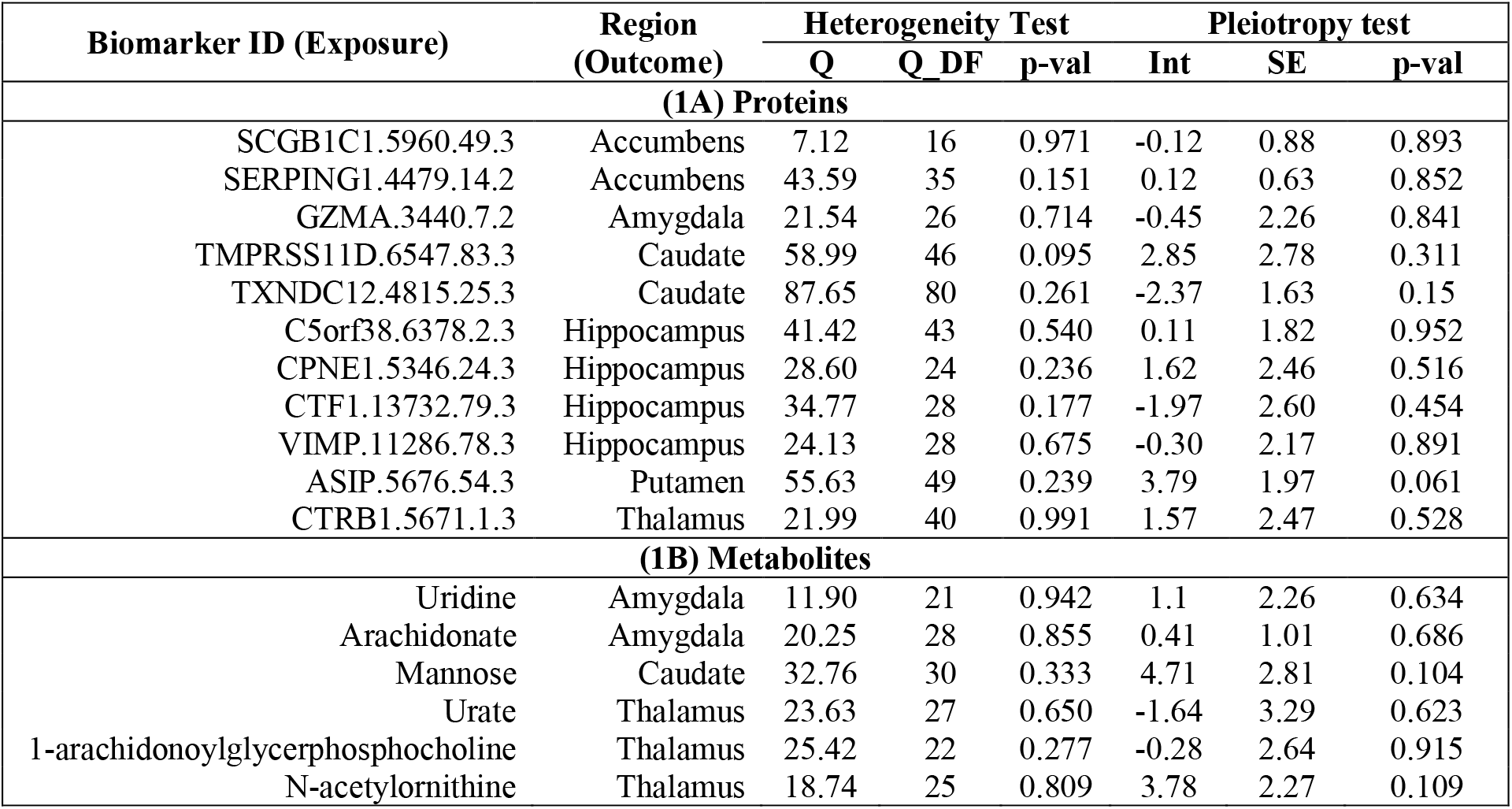
Statistical validation of MR results. The table shows the results of heterogeneity and pleiotropy tests performed for all biomarkers that had significant association with subcortical volume. Table 1A shows the results for proteins and 1B shows the results for the metabolites. Q refers to Cochran’s Q estimate for the heterogeneity test; DF is the degree of freedom. The Int refers to the MR-Egger intercept for the pleiotropy test and SE is the standard error of the Intercept.

### Reverse Mendelian Randomization Analysis

We performed the MR analysis with the subcortical brain structure volumes as exposure and the significantly associated biomarkers as outcomes. The results showed that for all proteins except C5orf38, there was no reverse causation observed in our analysis (**Table 2A**), thus indicating the causal effects of the proteins on the subcortical brain volume were statistically robust and not false positives. No reverse association was found between subcortical brain volume and the six metabolites as well (**Table 2B**).

**Table 2:**
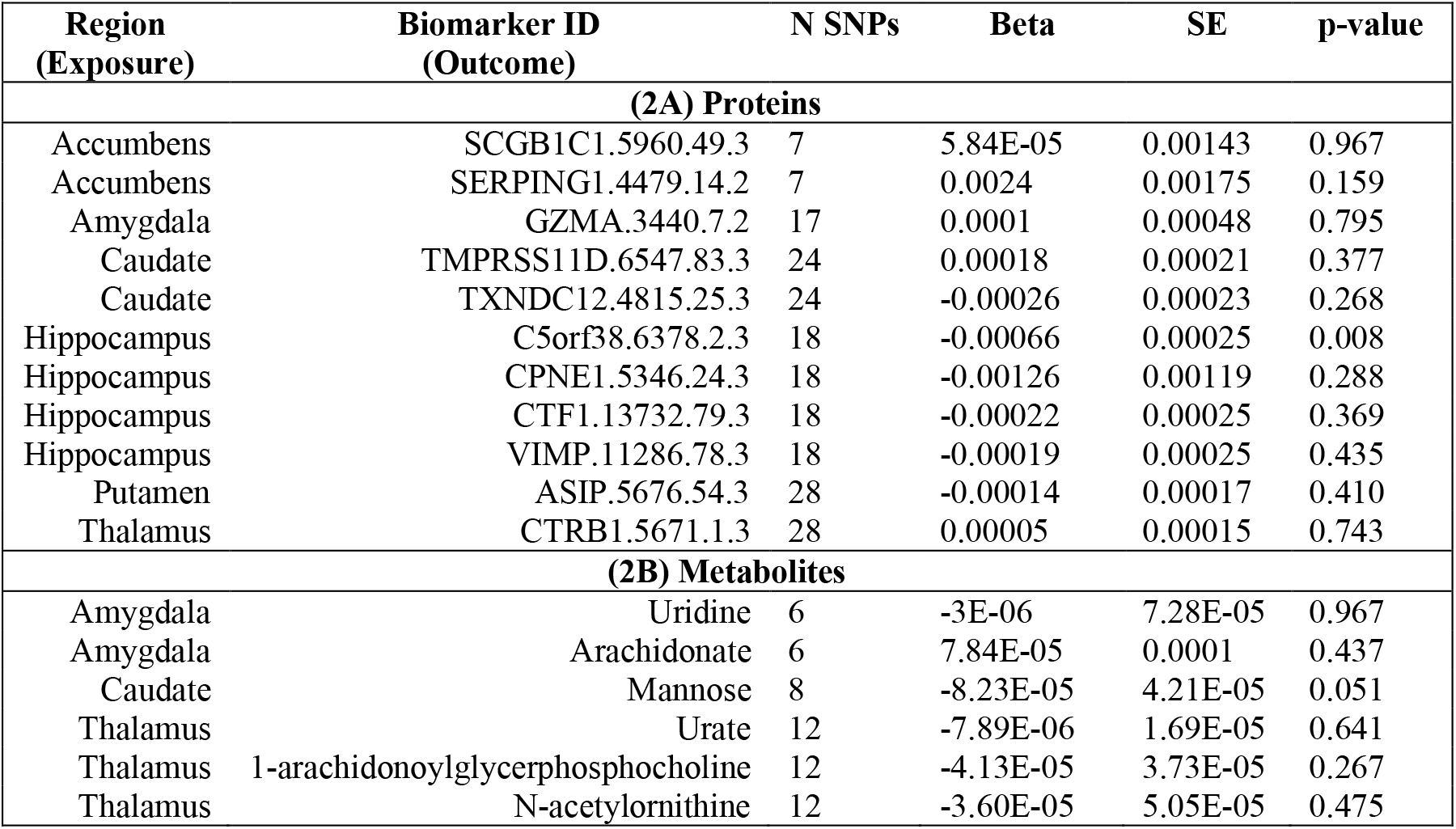
Reverse MR analysis. The table shows the results of MR analysis with the subcortical brain structures as exposure and the biomarkers that were significant in the primary analysis as the outcomes. Table 2A shows the results for proteins and 2B shows the results for the metabolites. N SNPs is the number of genetic instruments used for the analysis.

### Functional Enrichment Analysis

Analysis of the associated proteins using the g:Profiler platform revealed significant enrichment for various Gene Ontology terms after adjusting for multiple testing (**Figure 4** and **Supplementary Table 8**). These included molecular functions such as endopeptidase activity, peptidase activity and hydrolase activity. We also observed significant enrichment for biological processes such as proteolysis, regulation of the endoplasmic reticulum apoptotic signaling pathway and negative regulation of DNA binding. Most of the proteins were enriched in the extracellular regions of the human system. No significant enrichment was observed for the metabolites across all metabolic pathways.

**Figure 4:**
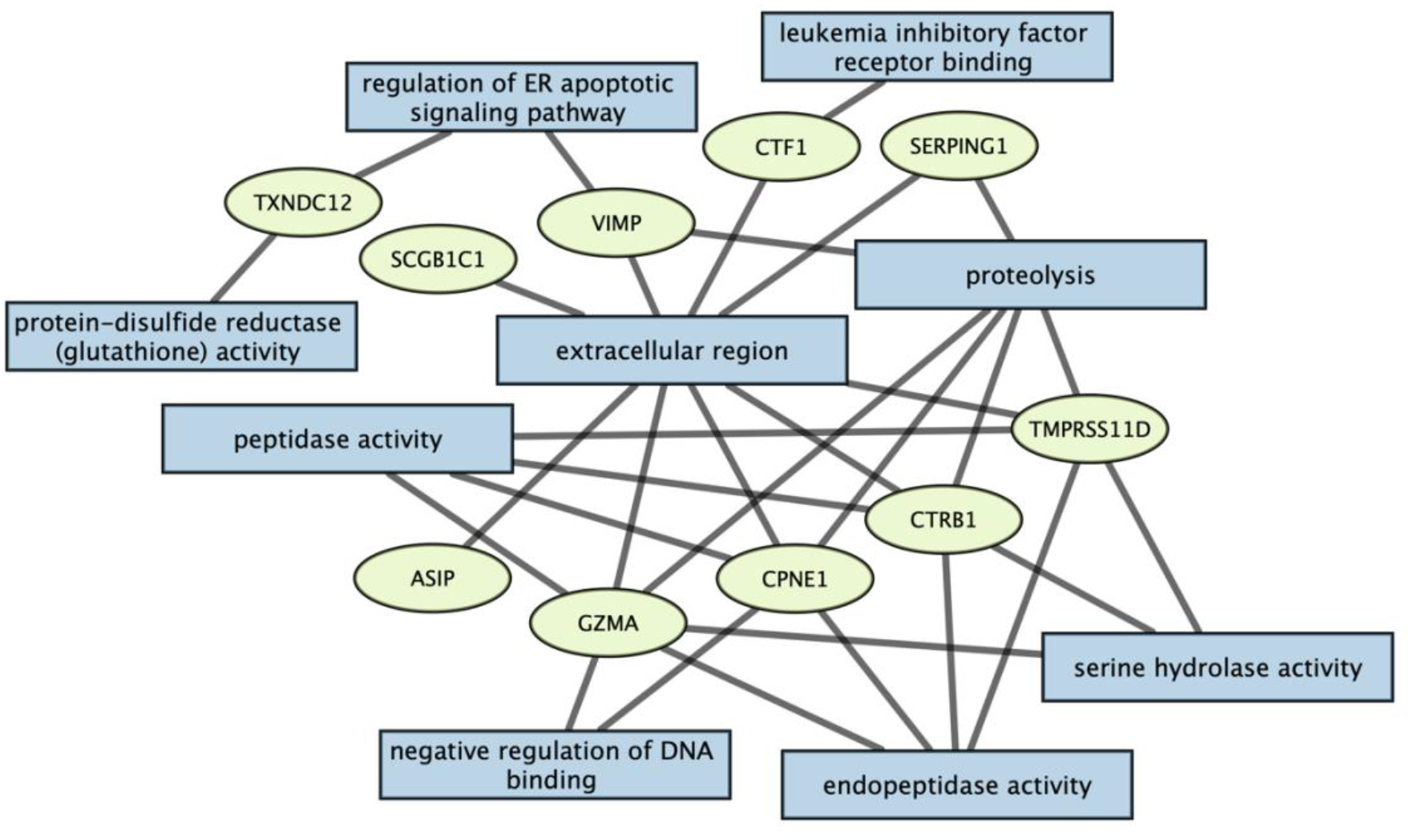
Enrichment analysis of proteins using the g:Profiler tool. The rectangles correspond to the various enriched Gene Ontology terms and the proteins associated with each term are shown in ellipses.

## Discussion

Here, pursuing a systems biology, multi-omic approach, we sought to provide insights into the mechanisms and mediators that underlie known associations of brain structures and neuropsychiatric disease. To do this, we performed a two-sample MR analysis to identify potentially causal associations between the genetically predicted levels of different biomarkers (plasma proteome, blood metabolome and gut microbiome) and the volumes of seven subcortical brain structures. Analyzing available summary statistics from large-scale GWAS, we identified eleven proteins and six metabolites to have a significant causal association with at least one subcortical structure after correcting for multiple testing. Heterogeneity and pleiotropy analysis showed low to no deviation from null thus validating our associations as truly significant. Bi-directional MR analysis for the significant associations showed no reverse causation for any proteins or metabolites except one (C5orf38, which is an unknown protein). Finally, enrichment analysis of the association proteins showed significant enrichment for proteolytic processes including endopeptidase, peptidase and hydrolase activities. No significant causal associations were observed between different bacterial genera in the gut microbiome and subcortical brain structures.

The molecular functions and the roles of the different proteins identified in this analysis as causally associated with subcortical brain volumes point to various pathways and mechanisms that could also help explain the relationship between subcortical structures and neuropsychiatric disorders. For example, SERPING1 - which is a Plasma Protease inhibitor - is a glycosylated protein involved in the regulation of the complement cascade and has been previously found to be associated with influencing frontal cortical thickness [57, 58]. The complement system itself has been implicated in depression, schizophrenia, and other neurodegenerative disorders as well [61], [62]. The nucleus accumbens has been an important brain region for regulating behaviors related to schizophrenia, depression and addiction [61, 62] and our results indicate that this regulation is being driven by levels of SERPING1, which is causally associated with accumbens volume. Similar relationships can also be observed for many of our identified proteins. GZMA, which is a serine protease involved in pyroptosis [65], is also found to have a lower expression in patients with major depressive disorder (MDD) compared to healthy controls [22]. Patients with MDD also tend to have decreased amygdala volume [23] which, based on our results, could be driven by GZMA. Another interesting example is that of TXNDC12, which is a member of the thioredoxin (Trx) superfamily. The Trx system is an antioxidant system that is important in maintaining sulfhydryl homeostasis protecting against oxidative stress [66]. Studies have pointed to the role of Trx-mediated oxidative stress in Parkinson’s disease-associated dopaminergic neuron degeneration, thus indicating that this protein might be an important regulator of the dopamine reward system [67], [68]. The caudate which is part of the striatum and connected to the substantia nigra is heavily involved in the reward system where the dopaminergic neurons are produced [69]. Changes in caudate volume have been found to be associated with disorders such as anorexia and Parkinson’s disease [70], [71] and our results point to TXNDC12 being the mediator of these associations via the dopaminergic neuron generation and reward system.

Some of the proteins we identified had an established role in brain development. For example, the three proteins we found causally associated with hippocampus volume were Copine-1, Cardiotrophin-1 and Selenoprotein. Copine 1 is a calcium dependent phospholipid binding protein and plays a role in neuronal progenitor cell differentiation and induces neurite outgrowth [72]. Similar to copine-1, cardiotrophin-1 is also involved in the differentiation of neuronal stem cells via a protein kinase dependent signaling pathway [73]. Selenoprotein is an important protein that mediates the levels of selenium in the brain and is important for mammalian brain development and other functions such as antioxidant protection and synaptic signaling [74].

Apart from these proteins, we also identified six metabolites that were causally associated with subcortical brain volume. Previous studies have shown that these metabolites have an important role in the functioning of the central nervous system and are also associated with different neurological disorders. Uric acid is considered a key antioxidant in humans [75], but high levels of this metabolite are associated with increased risk of disorders such as ASD and ADHD [76], [77]. Both of these disorders are also associated with reduced thalamic volume [78], [79], which could be explained by higher levels of uric acid based on our results. Interestingly, both uric acid and uridine are implicated in the development of Lesch-Nyhan syndrome which is a congenital disorder that affects brain structure and behavior of the affected individuals [80]. Other metabolites such as mannose and arachidonate which were identified in our study have also been found to be associated with disorders like anxiety and depression in mouse model systems [81]–[83].

There are certain limitations of this study. First, there were very few or no genome-wide significant SNPs to be used as instrument variables for many biomarkers in the MR analysis. To address this, we used a more exploratory threshold of 1e-05 for selecting genetic instruments, like previously done in multiple previous studies [40, 41, 48, 49]. We evaluated the strength for these genetic instruments using different statistical methods and found that they were valid for MR analysis. Second, the proteins and metabolites were quantified in the plasma for the GWAS analysis, which is a natural choice for biomarker-focused applications considering its convenience; however, we do not know whether these biomarkers would have had similar levels in specific brain regions, because of the existence of the blood-brain-barrier. To address this, we checked for the expression and presence of the different proteins and metabolites in the CNS. We found that most of them are highly expressed in different parts of the brain [84] and play an important role in its development and function (**Supplementary Table 9**). We would also like to point out that, we performed an MR study and identified several statistically causal risk factors associated with the subcortical brain volume, but these findings need further biological validation using experimental verification in cells and model systems. Based on statistical analysis, our study points to the most reliable targets for downstream investment, analysis and experimental validation and provides novel insights into the physiology of brain structures.

In conclusion, we identified several proteins and metabolites that are causally associated with the volume of subcortical brain structures. Our study highlighted the role of proteolytic and anti-oxidative components in the development and functioning of the brain. The biomarkers we identified could mediate the relationship between subcortical structures and different neurological and neuropsychiatric disorders. Future analysis could examine other characteristics of the brain such as neuronal activity, gray matter volume, and white matter connectivity which could further improve our understanding of the functioning of the central nervous system and its association to disease. The results of this study not only provide novel insight for understanding subcortical brain structure, but also help in uncovering potential diagnostic markers and drug targets for the many disorders that are associated with changes in brain structures.

## Data Availability

All data produced in the present work are contained in the manuscript

## Statement of Contribution

PJ, MY, CR, and PP designed the study, and performed primary analysis. All authors provided data, materials, and methods. All authors contributed to interpreting results, writing, reviewing, and editing the manuscript.

## Acknowledgments

This work was funded by NIH grants R01NS105746, R01MH126213 and NSF grants 1715202, and 2006929 awarded to Dr. Peristera Paschou.

## Data Availability

The GWAS summary statistics used for the analysis were downloaded from publicly available sources. The subcortical brain volume GWAS were downloaded from the ENIGMA consortium (https://enigma.ini.usc.edu/). The proteome GWAS were downloaded from the SomaLogic Plasma Protein GWAS study (http://www.phpc.cam.ac.uk/ceu/proteins/). The metabolites GWAS were obtained from Metabolites GWAS server (http://metabolomics.helmholtz-muenchen.de/gwas/) and the microbiome GWAS results were downloaded from https://mibiogen.gcc.rug.nl/.

