## Supplementary material for "Multiomic approach and Mendelian randomization analysis identify causal associations between blood biomarkers and subcortical brain structure volumes": Supplement MR.pdf

Jain, Yates et al.

7 MR Steiger Directionality Test – Metabolites……………………………………….. 8
8 Enrichment Analysis – Proteins…………………………………………………… 9
9 Protein and Metabolite Presence in brain……………………………………………. 10

**References**……………………………………………………………………………….11

**Supplementary Table 1**: GWAS studies for exposures and outcomes. Summary statistics were downloaded from publicly available data sources

| Dataset | N Features | GWAS Size | Ancestry | Source |
|---|---|---|---|---|
| Subcortical Brain Structures | Volume of 7 subcortical regions | 38,851 | European | [1] |
| Plasma Proteins | 3283 Somamers: 2994 Proteins | 3301 | European | [2] |
| Blood Metabolites | 237 metabolites | 7824 | European | [3] |
| Gut Microbiome | 103 Microbial Genera | 18,340 | Trans-Ancestry | [4] |

**Supplementary Table 2:** Plasma Proteome and Subcortical structure volume – Significant Associations. The table shows the proteins with significant association after multiple testing correction. The Exposure column refers to the Somamers that target the corresponding protein.

| Outcome | Exposure | Protein Target Name | Beta | P-value |
|---|---|---|---|---|
| Accumbens | SCGB1C1.5960.49.3 | Secretoglobin family 1C member 1 | 9.71 | 3.03E-05 |
| | SERPING1.4479.14.2 | Plasma protease C1 inhibitor | 6.37 | 6.96E-07 |
| Amygdala | GZMA.3440.7.2 | Granzyme A | 16.99 | 1.43E-05 |
| Caudate | TMPRSS11D.6547.83.3 | Transmembrane protease serine 11D | -26.80 | 7.06E-07 |
| | TXNDC12.4815.25.3 | Thioredoxin domain-containing protein 12 | 11.74 | 2.35E-06 |
| Hippocampus | C5orf38.6378.2.3 | Protein CEI | -21.24 | 9.82E-07 |
| | CPNE1.5346.24.3 | Copine-1 | -21.07 | 4.93E-05 |
| | CTF1.13732.79.3 | Cardiotrophin-1 | -25.94 | 8.92E-06 |
| | VIMP.11286.78.3 | Selenoprotein S | -25.54 | 4.26E-06 |
| Putamen | ASIP.5676.54.3 | Agouti-signaling protein | -27.99 | 1.21E-08 |
| Thalamus | CTRB1.5671.1.3 | Chymotrypsinogen B | -23.90 | 1.44E-05 |

**Supplementary Table 3:** Metabolites and Subcortical structure volume – Significant Associations. The table shows the metabolites with significant association after multiple testing correction. The Exposure column refers to the metabolite IDs.

| ***Outcome*** | **Exposure** | **Metabolite Name** | **Beta** | **Pval** |
|---|---|---|---|---|
| Amygdala | M00606 | Uridine | 255.95 | 1.44E-04 |
| | M01110 | Arachidonate | -110.44 | 1.83E-03 |
| Caudate | M00584 | Mannose | 244.78 | 5.54E-05 |
| Thalamus | M01604 | Urate | -458.7224 | 1.21E-05 |
| | M15630 | N-acetylornithine | 72.38 | 5.62E-04 |
| | M33228 | 1-arachidonoylglycerophosphocholine | 269.70 | 1.05E-04 |

**Supplementary Table 4:** Sensitivity Analysis – Proteins. The table shows results of sensitivity analysis between proteins (significant in primary analysis) and subcortical brain structure volume using two alternate methods of MR estimation.

| Outcome | Exposure | Method | N snps | Beta | SE | p-value |
|---|---|---|---|---|---|---|
| Accumbens | SCGB1C1.5960.49.3 | MR Egger | 17 | 10.30 | 4.91 | 0.05332 |
| | | Weighted median | 17 | 8.56 | 3.09 | 0.00563 |
| | SERPING1.4479.14.2 | MR Egger | 36 | 5.96 | 2.58 | 0.02726 |
| | | Weighted median | 36 | 4.24 | 1.83 | 0.02056 |
| Amygdala | GZMA.3440.7.2 | MR Egger | 27 | 20.20 | 13.57 | 0.14913 |
| | | Weighted median | 27 | 14.05 | 5.87 | 0.01667 |
| Caudate | TMPRSS11D.6547.83.3 | MR Egger | 47 | -37.43 | 11.71 | 0.00255 |
| | | Weighted median | 47 | -26.99 | 8.55 | 0.00160 |
| | TXNDC12.4815.25.3 | MR Egger | 81 | 16.16 | 3.92 | 0.00009 |
| | | Weighted median | 81 | 14.01 | 3.87 | 0.00030 |
| Hippocampus | C5orf38.6378.2.3 | MR Egger | 44 | -21.63 | 7.63 | 0.00701 |
| | | Weighted median | 44 | -23.97 | 6.65 | 0.00031 |
| | CPNE1.5346.24.3 | MR Egger | 25 | -25.31 | 8.30 | 0.00570 |
| | | Weighted median | 25 | -27.78 | 7.00 | 0.00007 |
| | CTF1.13732.79.3 | MR Egger | 29 | -19.39 | 10.44 | 0.07405 |
| | | Weighted median | 29 | -30.56 | 7.60 | 0.00006 |
| | VIMP.11286.78.3 | MR Egger | 29 | -24.53 | 9.18 | 0.01261 |
| | | Weighted median | 29 | -17.34 | 8.54 | 0.04229 |
| Putamen | ASIP.5676.54.3 | MR Egger | 50 | -39.49 | 7.66 | 0.00000 |
| | | Weighted median | 50 | -33.80 | 6.98 | 0.00000 |
| Thalamus | CTRB1.5671.1.3 | MR Egger | 41 | -28.91 | 9.81 | 0.00542 |
| | | Weighted median | 41 | -24.56 | 9.19 | 0.00750 |

**Supplementary Table 5:** Sensitivity Analysis – Metabolites. The table shows results of sensitivity analysis between Metabolites (significant in primary analysis) and subcortical brain structure volume using two alternate methods of MR estimation.

| Outcome | Exposure | Method | N snps | Beta | SE | pval |
|---|---|---|---|---|---|---|
| Amygdala | Uridine | MR Egger | 22 | 147.35 | 216.16 | 0.503 |
| | | Weighted median | 22 | 243.55 | 92.42 | 0.00841 |
| | Arachidonate | MR Egger | 29 | -111.91 | 54.42 | 0.0495 |
| | | Weighted median | 29 | -55.85 | 42.90 | 0.193 |
| Caudate | Mannose | MR Egger | 31 | 37.76 | 133.09 | 0.779 |
| | | Weighted median | 31 | 196.15 | 91.46 | 0.0320 |
| Thalamus | Urate | MR Egger | 28 | -433.75 | 231.35 | 0.0721 |
| | | Weighted median | 28 | -470.72 | 157.91 | 0.00287 |
| | N-acetylornithine | MR Egger | 26 | 40.65 | 28.33 | 0.164 |
| | | Weighted median | 26 | 57.82 | 25.82 | 0.025 |
| | 1-arachidonoylglycerophosphocholine | MR Egger | 23 | 20.01 | 24.03 | 0.414 |
| | | Weighted median | 23 | 15.93 | 17.96 | 0.375 |

**Supplementary Table 6:** Directionality Test – Proteins. The table shows results of directionality test between proteins (significant in primary analysis) and subcortical brain structure volume using MR Steiger test. The r2 refers to the variance explained by the SNP instrument in the exposure and outcome respectively.

| Outcome | Exposure | snp_r2 (exposure) | snp_r2 (outcome) | Correct causal direction | Steiger p-value |
|---|---|---|---|---|---|
| Accumbens | SCGB1C1.5960.49.3 | 0.1358 | 0.0022 | TRUE | 5.04E-66 |
| | SERPING1.4479.14.2 | 0.5048 | 0.0058 | TRUE | 0 |
| Amygdala | GZMA.3440.7.2 | 0.1973 | 0.0034 | TRUE | 1.33E-100 |
| Caudate | TMPRSS11D.6547.83.3 | 0.5856 | 0.0074 | TRUE | 0 |
| | TXNDC12.4815.25.3 | 2.5944 | 0.0096 | TRUE | 0 |
| Hippocampus | C5orf38.6378.2.3 | 0.6847 | 0.0055 | TRUE | 0 |
| | CPNE1.5346.24.3 | 0.5622 | 0.0045 | TRUE | 0 |
| | CTF1.13732.79.3 | 0.4957 | 0.0051 | TRUE | 0 |
| | VIMP.11286.78.3 | 0.4398 | 0.0038 | TRUE | 3.77E-303 |
| Putamen | ASIP.5676.54.3 | 1.0082 | 0.0079 | TRUE | 0 |
| Thalamus | CTRB1.5671.1.3 | 0.6757 | 0.0033 | TRUE | 0 |

**Supplementary Table 7:** Directionality Test – Metabolites. The table shows results of directionality test between Metabolites (significant in primary analysis) and subcortical brain structure volume using MR Steiger test. The r2 refers to the variance explained by the SNP instrument in the exposure and outcome respectively.

| Outcome | Exposure | snp_r2 (exposure) | snp_r2 (outcome) | Correct causal direction | Steiger p-value |
|---|---|---|---|---|---|
| Amygdala | Uridine | 0.0851 | 0.0021 | TRUE | 3.66E-66 |
| | Arachidonate | 0.1782 | 0.0026 | TRUE | 4.28E-155 |
| Caudate | Mannose | 0.1989 | 0.0041 | TRUE | 3.91E-165 |
| Thalamus | Urate | 0.2108 | 0.0034 | TRUE | 6.24E-192 |
| | N-acetylornithine | 0.3530 | 0.0024 | TRUE | 0 |
| | 1-arachidonoylglycerophosphocholine | 0.1495 | 0.0021 | TRUE | 3.377E-128 |

**Supplementary Table 8:** Enrichment analysis - Proteins. The table shows results of enrichment analysis using g:Profiler platform. GO: Gene ontology, MF: Molecular Function, BP: Biological Processes, CC: Cellular Components.

| Source | Term Name | p-val (adj) | Protein Intersections |
|---|---|---|---|
| **GO:MF** | endopeptidase activity | 0.003 | P12544, O60235, Q99829, P17538 |
| **GO:MF** | serine-type endopeptidase activity | 0.005 | P12544, O60235, P17538 |
| **GO:MF** | serine-type peptidase activity | 0.006 | P12544, O60235, P17538 |
| **GO:MF** | serine hydrolase activity | 0.007 | P12544, O60235, P17538 |
| **GO:MF** | peptidase activity | 0.011 | P12544, O60235, Q99829, P17538 |
| **GO:MF** | protein-disulfide reductase (glutathione) activity | 0.031 | O95881 |
| **GO:MF** | leukemia inhibitory factor receptor binding | 0.043 | Q16619 |
| **GO:BP** | proteolysis | 0.030 | P05155, P12544, O60235, Q99829, Q9BQE4, P17538 |
| **GO:BP** | regulation of endoplasmic reticulum stress-induced intrinsic apoptotic signaling pathway | 0.035 | O9588, Q9BQE4 |
| **GO:BP** | negative regulation of DNA binding | 0.048 | P12544, Q99829 |
| **GO:CC** | extracellular region | 0.00001 | Q8TD33, P05155, P12544, O60235, Q99829, Q16619, Q9BQE4, P42127, P17538 |
| **GO:CC** | extracellular space | 0.016 | P05155, O60235, Q99829, Q16619, Q9BQE4, P42127, P17538 |

**Supplementary Table 9:** Protein and Metabolite Presence in brain. The table shows whether the biomarkers identified are expressed in brain or can cross the blood brain barrier (BBB) and their functions

| Biomarker ID (Outcome) | Present in Brain (Yes/No/NA) | Function |
|---|---|---|
| SCGB1C1 | Yes | Primary host defense mechanism [5] |
| SERPING1 | Yes | Cortical Development [6] |
| GZMA | Yes | Pyroptosis related cell death [7] |
| TMPRSS11D | Yes | Host Defense System [8] |
| TXNDC12 | Yes | Protection against oxidative stress [9] |
| C5orf38 | NA | - |
| CPNE1 | Yes | Neuronal progenitor cell differentiation [10] |
| CTF1 | Yes | Neuronal stem cell differentiation [11] |
| VIMP | Yes | Synaptic signaling and brain development [12] |
| ASIP | No | Regulation of melanogenesis [13] |
| CTRB1 | No | Precursor or proteolysis enzyme [14] |
| Uridine | Yes | Neuronal membrane formation [15] |
| Arachidonate | Yes | Functioning of ion channels [16] |
| Mannose | Yes | Promotes anti-inflammatory response [17] |
| Urate | Yes | Protection from oxidative damage [18] |
| 1-arachidonoylglycerphosphocholine | NA | - |
| N-acetylornithine | Yes | Neuronal regulation and axon signaling [19] |
